# Correlation between Galanin and its receptor with the serum electrolytes in Long-COVID patients

**DOI:** 10.1101/2023.11.27.23299076

**Authors:** Wasim Talib Mahdi Al Masoodi, Sami Waheed Radhi, Habiba Khdair Abdalsada, Hussein Kadhem Al-Hakeim

## Abstract

**Background:** Long-COVID is a complicated condition with prolonged SARS-CoV-2 symptoms. Several variables have been studied in this illness. Among the less studied variables are galanin and its receptor (GalR1). The Galanin system is involved in the pathophysiology of several age-related chronic disorders, including alcoholism, chronic pain, and bowel and skin inflammation. The aim of the study is to correlate the galanin system parameters with clinical and biochemical variables in Long-COVID. **Methods:** Serum levels of albumin, electrolytes, GAL, GALR1, and C-reactive protein (CRP) are measured by ELISA technique in 90 Long-COVID patients and 60 recovered subjects who are free from any symptoms of Long-COVID. **Results:** The study showed a significantly increased Galanin, GALR1, and the Gal/GALR1 ratio. On the contrary, serum albumin, total calcium, ionized calcium, total magnesium, and the ionized calcium/magnesium ratio were significantly decreased. Galanin and Galanin/GALR1 showed significant age-related associations (ρ=0.353, p<0.01) and (ρ=0.218, p<0.05), respectively. The lowest SpO2 was associated with Galanin (ρ=-0.295, p<0.01) and GALR1 (ρ=-0.232, p<0.05), respectively. According to ROC analysis results, the highest sensitivities for differentiating between patients and non-patient subjects were Galanin (71.7%) and GALR1 (60.0%). **Conclusions:** Galanin, GALR1, and Long-COVID disease are directly correlated. However, more research is needed to find out exactly what roles plasma Galanin and its receptor play in Long-COVID disease.

## Introduction

Corona virus disease-2019 (COVID-19) is a complex systemic disease, affecting the cardiovascular, renal, hematologic, gastrointestinal and central nervous systems (CNS) [1], with a significant global morbidity and mortality [2]. COVID-19 causes multi-organ impairments with a significant number of survivors experiencing Long-COVID Syndrome [3]. About 30%–60% of asymptomatic or mild COVID-19 people acquire a Long-COVID [4]. The WHO defines Long-COVID as a post-COVID-19 condition that “occurs in individuals with a history of probable or confirmed SARS-CoV-2 infection, usually 3 months from COVID-19 onset with various symptoms, even weeks or months after acquiring it irrespective of viral status” [5, 6]. Long-COVID has complex, varied symptoms due to immune system problems [7, 8]. Long-COVID research, diagnosis, and treatment should be defined and harmonized to improve national and international data collecting and age-specific incidence, prevalence, and risk factors [9]. Long-COVID may not be caused by SARS-CoV-2, but by COVID-19’s biopsychosocial consequences[10]. Long-term COVID can impact adults, children, and adolescents, and many did not return to their past work and quality of life [11]. Globally, Long-COVID-19 can infect anyone with SARS-CoV-2, independent of acute illness severity [12, 13]. Long-COVID can cause a variety of new or persistent symptoms over weeks or months [14]. After symptoms appear, recuperation takes 7-10 days, although severe illness can take two to 12 months [5, 15–17]. The symptoms are varied and organ-related, including neurological, thoracic, stomach, ears, nose, throat, eyes, vascular, hair, skin, and genitourinary [18]. These symptoms include persistent fevers, digestive issues, blood and mucosa abnormalities, and presumably greater organ-targeted autoantibodies [11, 19, 20]. Some persons have tachycardia, intense exhaustion, and incapacity to perform daily physical duties [21]. Long-COVID is a complex illness with long-term diverse symptoms, requiring special treatment and continuing assistance [22]. In order to detect and monitor Long-COVID pathology, neuronal injury biomarkers and electrolyte molecules have been studied. Plasma Galanin (GAL) and GALR1 have not been studied in Long-COVID patients. Plasma GAL and GALR1 alterations may also be connected to Long-COVID symptoms. This is a knowledge gap, as their roles in Long-COVID have not been widely studied, despite being primarily associated with nervous system functions and various physiological roles. Current study focuses on galanin, GALR1, and Long-COVID. Galanin is a neuropeptide encoded by the GAL gene [23]. It modulates immune responses via three G-protein coupled receptors [36]. Its highest affinity for GALR1 and GALR2 [24], released by the enteric nervous system neurons that act by binding to GAL receptors 1, 2, and 3 (GALR1, 2, and 3), can also modulate depression, anxiety, pain threshold, and pain behaviors [25, 26]. Galanin inhibits myenteric neuron transmitter release via GALR1 [27]. High expression of GALR1 was observed in GABA neurons [28]. Galanin, its receptor, and GAL/GAL1R went down [29]. The liver produces albumin, the primary protein in blood, which regulates blood oncotic pressure, binds water, cations, fatty acids, hormones, bilirubin, T4, and medicines [30, 31]. Albumin serves as a major anti-inflammatory agent in our body [32]. COVID-19 patients with low serum albumin have worse results, including death [33], and predicting COVID-19 severity [34]. Serious COVID-19 is linked to serum electrolyte abnormalities [35, 36]. Drug side effects, fever, hyperventilation, perspiration, and nutrition may induce it [37]. After treating severe COVID-19, calcium insufficiency may improve [38]. Non-specific C-reactive protein (CRP) indicates infection, inflammation, and tissue injury [39]. Concentration indicates disease severity [40]. CRP was increased in Long-COVID patients [41]. Higher stigma levels were linked to COVID-19 survivor status, economic loss, and depressed symptoms [42]. The study evaluated neural injury markers, serum electrolytes, and blood markers in individuals with prolonged COVID progression, using ROC analysis to predict severe outcomes, including sociodemographic and electrolytes.

## Subjects and Methods

### Participants

The present case-control study involved 90 patients with long-COVID-like symptoms who had been diagnosed and treated for acute COVID-19 and 60 controls who did not develop Long-COVID in the first three months of 2023. The patients were identified based on the WHO post-COVID (Long-COVID) rules [43], which include the following: (a) individuals with a history of proven SARS-CoV-2 infection; (b) symptoms that persisted past the acute stage of illness or manifested during recovery from acute COVID-19 infection; (c) symptoms that lasted at least 2 months and are present 3–10 months after the onset of COVID-19; furthermore, (d) patients who have no less than two symptoms that make it challenging to do everyday undertakings, for example, fatigue, headaches, trouble speaking, chest pain, a persistent cough, loss of taste or smell, emotional symptoms, mental hindrance, or fever [43]. None of these criteria were met by the 36 controls. All individuals diagnosed with acute COVID-19 received treatment at a designated quarantine hospital located in Kerbala City. These hospitals include Imam Al-Hussein Medical City of Kerbala, Imam Al-Hassan Al-Mujtaba Teaching Hospital, Karbala Teaching Hospital for Children, Alkafeel Super Specialty Hospital, and Al-Hindiyah General Hospital. Based on the presence of typical symptoms like fever, breathing issues, coughing, and loss of smell and taste, positive reverse transcription real-time polymerase chain reaction findings, and positive IgM directed to SARS-CoV-2, senior physicians and virologists diagnosed SARS-CoV-2 infection and acute COVID-19. The patients’ rRT-PCR results following the acute phase were all negative. We selected 36 controls, their family members, or their companions from a similarly exclusive location. Additionally, we included controls who tested negative for rRT-PCR but did not exhibit any clinical symptoms of acute phase disease, such as a dry cough, sore throat, fatigue, lack of appetite, influenza-like symptoms, fever, night sweats, or chills. Furthermore, the study excluded individuals with diabetes mellitus or other systemic autoimmune diseases, multiple sclerosis, stroke, neurodegenerative or neuroinflammatory disorders, psoriasis, rheumatoid arthritis, inflammatory bowel disease, scleroderma, liver disease, or any other medical conditions. We recorded Sinopharm, Pfizer, and AstraZeneca vaccinations. Body mass index (BMI) was computed by dividing body weight (kg) by height in meters squared. Additionally, the study did not include pregnant or breastfeeding women. Before participating in the study, all control and patient participants, or their respective parents or legal guardians, provided written consent after receiving comprehensive information. The Kerbala Health Directorate-Training and Human Development Centre (Document No.0030/2023) and the University of Kufa institutional ethics committee (1657/2023) approved the study. The study adhered to both Iraqi and international ethical and privacy laws, including the International Conference on Harmonization of Good Clinical Practice, the Belmont Report, the CIOMS Guidelines, and the World Medical Association’s Declaration of Helsinki.

### Assays

After awakening and before consuming breakfast, early morning blood samples were taken around 9 a.m. A volume of five milliliters of venous blood was obtained and transferred into plain, sterile tubes. Any samples that were hemolyzed were excluded from further analysis. The blood samples that had formed clots were subjected to centrifugation at a speed of 3,000 rpm for a duration of five minutes after ten minutes had elapsed. Subsequently, the serum was separated and transferred into three new Eppendorf tubes to be stored at −80 °C until thawed for assay. The ELISA technique was employed to quantify serum human Galanin and GALR1 by ELISA kits supplied by Nanjing Pars Biochem Co., Ltd. (Nanjing, China). The Gal and GALAR1 tests exhibited a sensitivity range of 10 ng/L to 300 ng/L, with an intra-assay coefficient of variation (CV) of 10.0% (precision within-assay). CRP levels in human serum were assessed using CRP latex slide assays (Spinreact®, Barcelona, Spain). The levels of calcium, magnesium, albumin, and glucose in serum were determined spectrophotometrically using a ready-to-use kit provided by Spinreact® (Barcelona, Spain). We strictly adhered to the manufacturer’s instructions, following each step precisely.

### Statistical analysis

The Kolmogorov-Smirnov test analysed findings group distributions. By statistical distribution, nonparametric and regularly distributed variable findings are distinguished. (Mean Standard Deviation) was used to express the results for the normally distributed variable. Results were presented for nonparametric variables such medians and interquartile ranges of 25%–75%. The Mann-Whitney U test was used to compare the subdivided groups in the measurement parameters between the control groups and those suffering groups. Calculating the correlation Spearman’s coefficients (rho) yields an approximation of the degree of the parameters’ correlation. Analysis of variance (ANOVA) was used to look at differences in continuous variables between groups and contingency tables (χ ^2^-test) to look at associations between nominal variables. The two variables’ relationship was examined using Pearson’s product-moment correlation coefficients. To assess the diagnostic utility of the detected biomarkers, receiver operating characteristics (ROC) analysis was utilized. Calculated variables include cut-off points, sensitivities, specificities, and Youdin’s statistic. For all statistical calculations, SPSS version 26 software’s two-tailed tests with a p-value cutoff of 0.05 were employed. Using Excel 2019 from Microsoft Office, the figures were organized.

## Results

### Sociodemographic and clinical data

In Table 1, demographic and clinical data from acute COVID-19 patients without symptoms (No Long-COVID) and those with Long-COVID are shown. Long-COVID patients and healthy controls demographics (age, sex, height, weight, BMI, residency, marital status ratio, smoker, vaccination with A1, PF2, S3, and paracetamol, azithromycin, VitC, VitD, and Zn treatment) were not significantly different. At the same time, it is expected to find a significant decrease in the duration of disease (p = 0.001), highest body temperature (p = 0.005), education (p = 0.004), employment (p = 0.002), duration of disease days, period of cure months (p = <0.001), taking ceftriaxone (p = <0.001), dexamethasone (p = 0.004), and a decrease in the SpO2 (p = 0.001) in the patients group during the acute phase of infection with SARS-CoV-2 than the control group. Socioeconomic disparities, ignorance, occupational attributes, health literacy, and healthcare access increase acute SARS-CoV-2 infection in low-educated individuals. Variations in virus load, immunological response, treatment methods, and host features impact disease severity. Dexamethasone affects SARS-CoV-2 and acute COVID-19 differently.

**Table 1:**
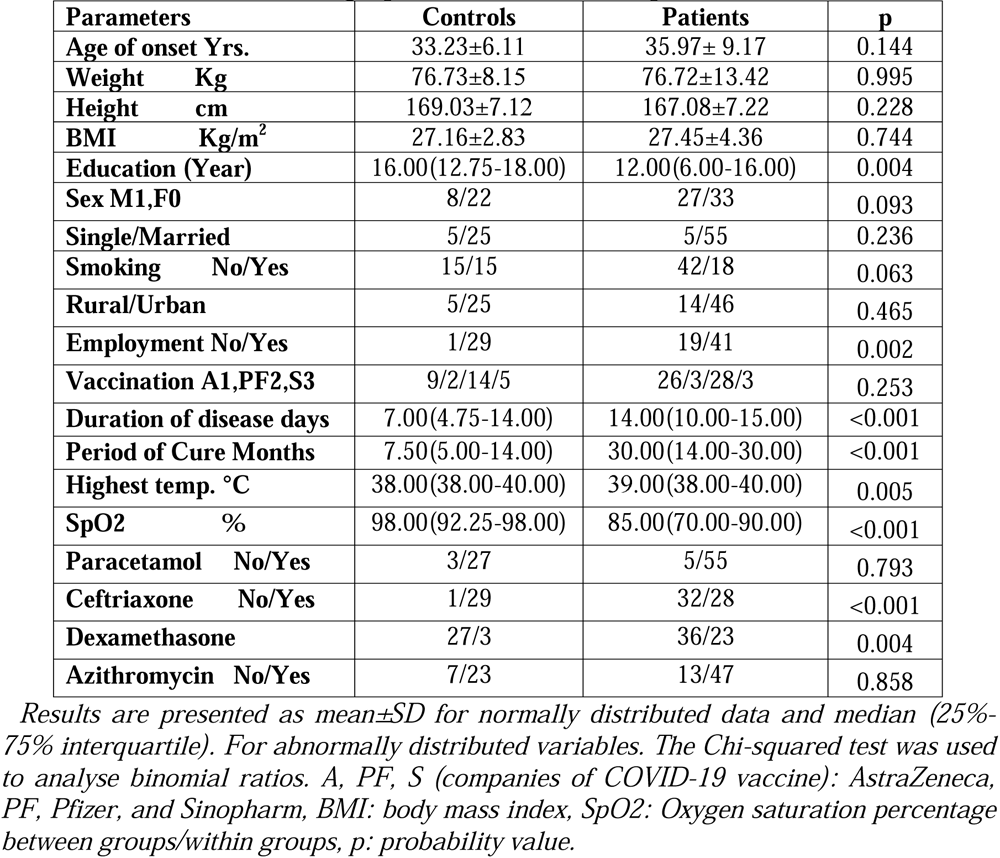
Socio-demographic characteristics of patients and control.

### Levels of serum albumin and electrolyte levels in Long-COVID and controls

Table 2 displays the findings about the serum albumin and electrolyte levels in individuals affected by Long-COVID and those who are considered healthy. The findings of the study indicate that individuals with Long-COVID had a statistically significant decrease in serum albumin levels (p = 0.300) compared to the control group of healthy individuals (HC). Additionally, Long-COVID patients demonstrated decreased levels of total calcium (T.Ca) (p = 0.082), ionized calcium (p = 0.061), total magnesium (T. Mg) (p = 0.056), ionized magnesium, total calcium to magnesium ratio (T.Ca/Mg) (p = 0.254), and ionized calcium to magnesium ratio (p=0.488). There is no statistically significant difference between the groups.

**Table 2:** The results of the serum albumin and electrolytes levels in LC patients and HC control groups.

| Parameters | Controls | Patients | $F/\chi^2$ | p |
| --- | --- | --- | --- | --- |
| Albumin g/l | 45.581±1.901 | 46.282±3.392 | 1.089 | 0.300 |
| T.Ca mM | 2.336±0.242 | 2.235±0.263 | 3.104 | 0.082 |
| I.Ca | 1.301±0.065 | 1.274±0.065 | 3.592 | 0.061 |
| T. Mg mM | 1.061±0.345 | 0.898±0.392 | 3.765 | 0.056 |
| I. Mg mM | 0.739±0.228 | 0.632±0.259 | 3.765 | 0.056 |
| T.Ca/Mg | 2.543±1.188 | 1.009±7.244 | 1.319 | 0.254 |
| I.Ca/Mg | 1.992±0.836 | 4.708±21.282 | 0.486 | 0.488 |

T. Ca: total calcium, I.Ca: ionized Calcium, Mg: Magnesium, $F/\chi^2$ : F-statistics value for continuous variables or Chi-square statistic value for categorical variables, df: degree of freedom between groups/within groups, p: probability value.

### Galanin and its receptor

Figure 1 displays the serum Gal (pg/mL) levels observed in both Long-COVID patients and the healthy controls (HC) groups. The findings of the study revealed a statistically significant rise (p<0.001) in Galanin levels among individuals diagnosed with Long-COVID [63.211 (51.577-141.616)] pg/ml in comparison to the control group [47.350 (41.367-57.084)] pg/ml. Figure 2 displays the serum GALR1 results for both the HC and Long-COVID (LC) groups. The findings of the study indicated a statistically significant elevation (p = 0.007) in GALR1 levels among individuals diagnosed with Long-COVID [1.965 (1.661-2.547)] pg/ml, in comparison to the HC group [1.770 (1.487-1.878)] pg/ml. Figure 3 displays the outcomes of the Gal/GalR1 assay conducted on the serum samples obtained from patients diagnosed with Long-COVID and a control group of individuals without the condition. The findings of the study revealed a statistically significant rise (p=0.019) in the Gal/GalR1 levels among individuals diagnosed with Long-COVID [32.957 (26.627-60.056)] in comparison to the HC group [26.741 (22.650-35.396)].

**Figure 1.**
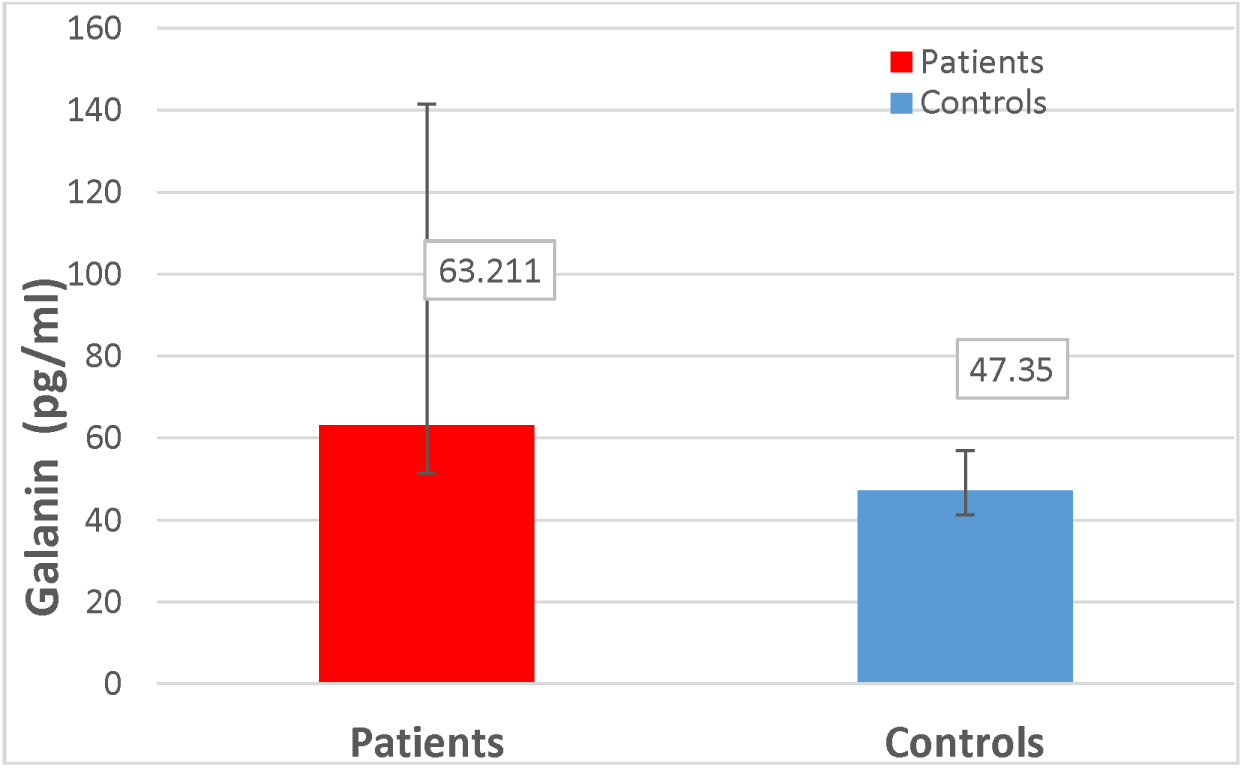
Serum Galanin in patients and control groups.

**Figure 2.**
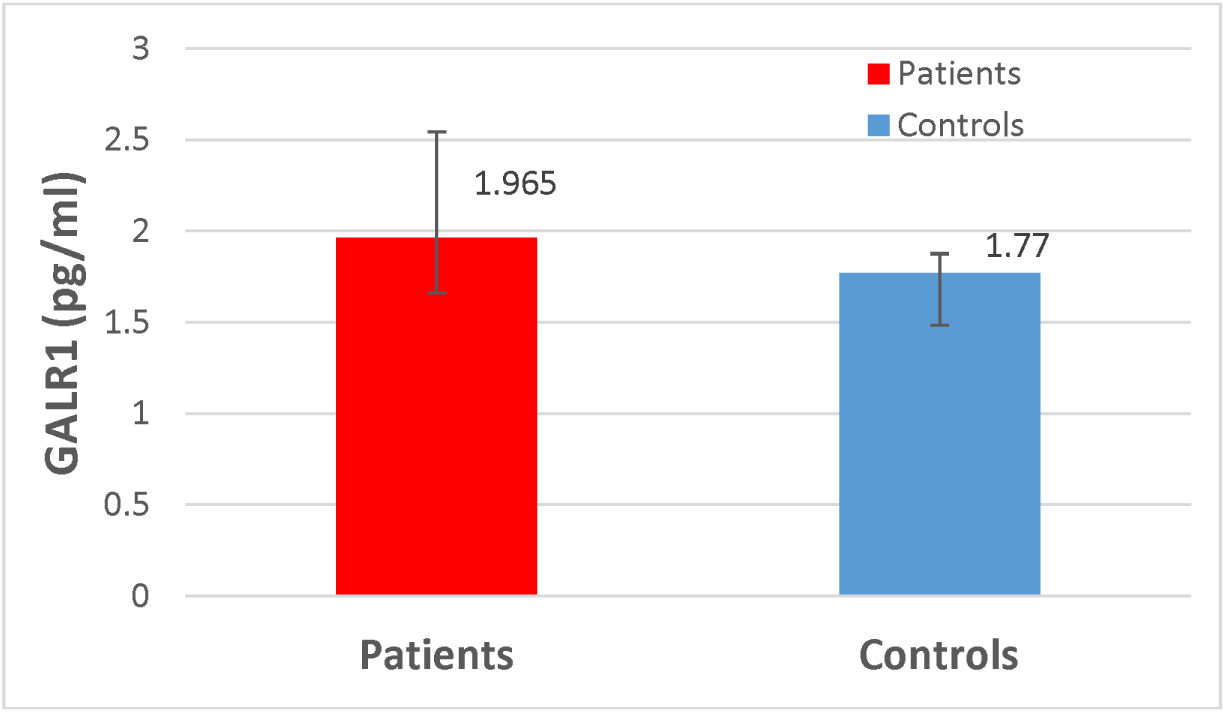
Serum GALR1 in patients and control groups.

**Figure 3.**
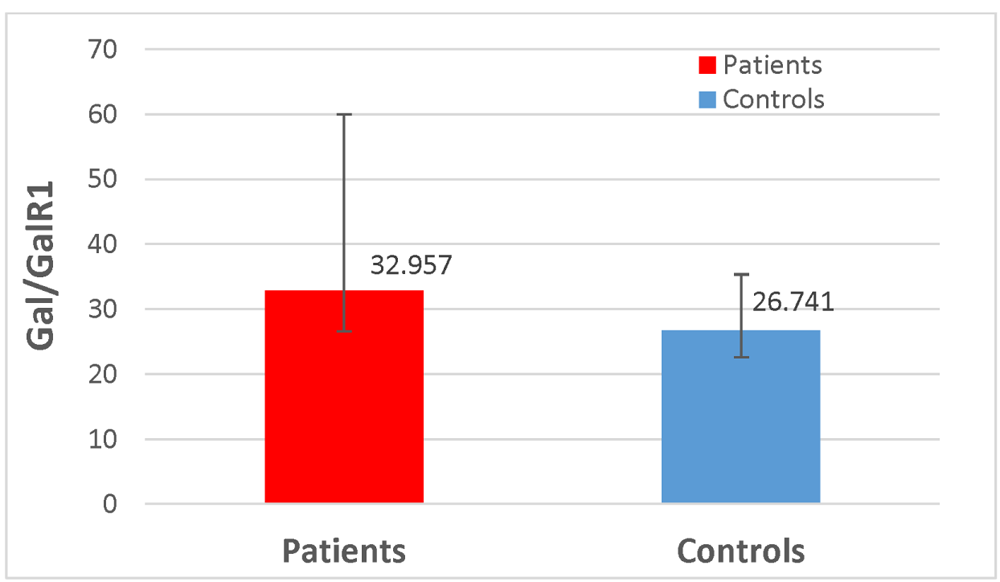
Serum Gal/GalR1 in patients and control groups.

### Intercorrelation matrix

Table 3 displays the findings of the link between the measured biomarkers and the sociodemographic information in the Long-COVID group. The findings demonstrated a statistically significant positive association between Galanin levels and age (ρ = 0.353, p<0.01). The findings indicated that there was a negative association (ρ = -0.295, p<0.01) between galanin and SpO2. The findings revealed a statistically significant negative connection (ρ = -0.232, p<0.05) between GALR1 and SpO2. The findings of the study revealed a statistically significant positive association between the levels of Gal/GalR1 and age (ρ = 0.218, p<0.05). Although sociodemographic data did not exhibit a significant association with other biomarkers.

**Table 3:** Correlations between biomarkers and socio-demographic characteristics in patients.

|  | Galanin | GALR1 | Gal/GalR1 |
| --- | --- | --- | --- |
| Sex | 0.013 | -0.021 | 0.075 |
| Age | 0.353** | 0.124 | 0.218* |
| BMI | 0.153 | -0.002 | 0.143 |
| Mar. State | 0.106 | 0.053 | 0.093 |
| Education years | -0.042 | -0.144 | 0.086 |
| Vaccination | 0.051 | -0.125 | 0.132 |
| Smoking | -0.009 | 0.169 | -0.099 |
| Residency | -0.051 | -0.139 | -0.003 |
| Employment | -0.066 | -0.072 | -0.018 |
| Duration of Dis. | -0.006 | -0.020 | 0.064 |
| Period of Cure | 0.048 | 0.130 | -0.022 |
| Highest temp. | 0.201 | 0.134 | 0.048 |
| SpO2 | -0.295** | -0.232* | -0.106 |

\* Correlation is significant at the 0.05 level (2-tailed).
\*\* Correlation is significant at the 0.01 level (2-tailed).

Table 4 displays the findings of the association between the cation levels in the LC group and the measured biomarkers. The findings of the study revealed a statistically significant positive association between Galanin and GalR1 (ρ=0.212, p<0.05), as well as between Gal/GalR1 (ρ=0.641, p<0.01). The findings of the study demonstrated a statistically significant link between GALR1 and Galanin (ρ=0.212, p<0.05), as well as an inverse correlation with Gal/GalR1 (ρ=-0.489, p<0.01). In contrast, the analysis revealed that there was no statistically significant association between cations and other biomarkers.

**Table 4:**
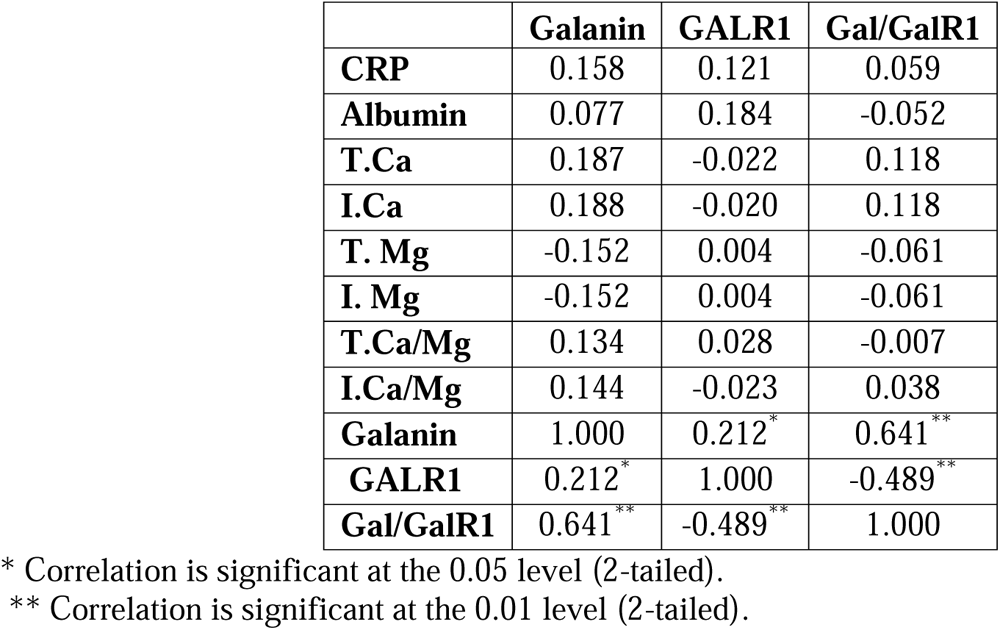
Correlations between among biomarkers and cations levels characteristics in patients.

### Receiver operating characteristic (ROC) analysis of biomarker

The application of receiver operating characteristic (ROC) analysis was employed to determine the diagnostic sensitivity and specificity of Galanin, GALR1, and the Gal/GalR1 ratio in the context of Long-COVID. The ROC curves are depicted in Figure 4 for analysis. Table 5 displays the optimal sensitivities and specificities achieved by varying the ROC coordinates and concentration cut-off. The findings shown in Table 5 and Figure 4 indicate that Galanin and GALR1 exhibit the highest sensitivities, specifically 71.7% and 60.0% respectively, in discerning individuals with Long-COVID from those who are healthy. Although the diagnostic significance of Gal/GalR1 is not statistically significant (p > 0.05), the findings shown in the Table can be interpreted in the following manner: Subjects exhibiting Galanin (Pg/ml) levels surpassing the established threshold value of 53.302 Pg/ml may potentially indicate the presence of LC, with a sensitivity of 71.7% and a specificity of 73.3%. Long-COVID patients may exhibit elevated levels of GALR1 (pg/ml) beyond the established cut-off value of 1.815 pg/ml, with a sensitivity and specificity of 60.0%. When the GalR1/Gal level exceeds the established threshold of 30.740 pg/ml, it indicates that individuals with Long-COVID have a sensitivity of 58.3% and a specificity of 57.6%.

**Figure 4.**
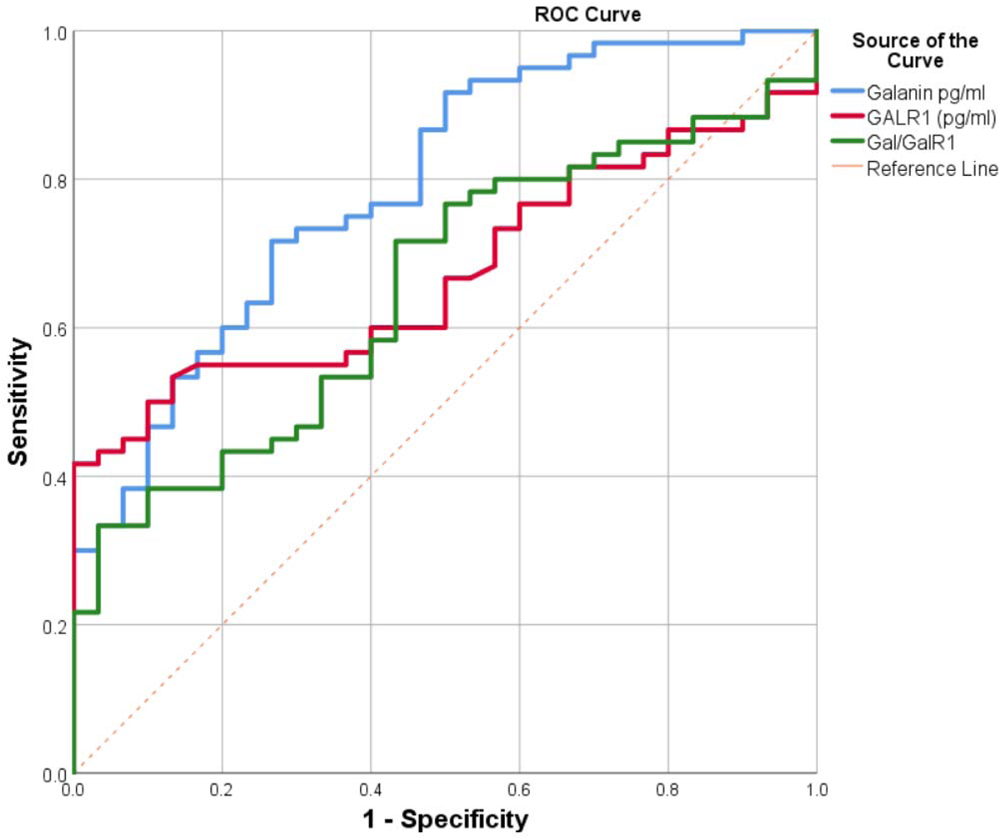
Receiver operating characteristic curves of Galanin, GALR1 and Gal/GalR1 in LC group.

**Table 5.** AUC study of receiver operating characteristics of Galanin, GALR1 and Gal/GalR1 in LC. group. CI: Confidence interval.

| **Variable(s)** | **Cut-Off** | **Sensitivity** | **Specificity** | **AUC(SE)** | **p-value** |
| --- | --- | --- | --- | --- | --- |
| **Galanin Pg/ml** | 53.302 | 71.7 | 73.3 | 0.790(0.049) | <0.001 |
| **GALR1 (Pg/ml)** | 1.815 | 60.0 | 60.0 | 0.674(0.055) | 0.005 |
| **Gal/ GalR1** | 30.740 | 58.3 | 57.6 | 0.652(0.058) | 0.713 |

## Discussion

Serum albumin levels in Long-COVID patients are lower than controls which was found previously [44]. Serum albumin had a substantial negative connection with serum albumin levels in severe COVID-19 patients, which may continue post-recovery [44]. COVID-19 infection with low serum albumin has significant consequences, including death [45]. Decreased serum albumin levels are linked to COVID-19 severity and poor outcomes [33]. For Long-COVID patients, long-term sickness, liver dysfunction, gastrointestinal difficulties that restrict nutrient absorption, and persistent inflammation may explain the substantial decline in serum albumin relative to the control group. Different physiological effects of Long-COVID are shown here. Compared to HC controls, Long-COVID patients exhibited significantly decreased T.Ca, ionized Ca, T.Mg, ionized Mg, T.Ca/Mg, and ionized Ca/Mg ratios. Studies demonstrate no significant difference across groups, supporting these findings: Serum calcium is lower in COVID-19 patients than in controls [46]. COVID-19 patients had significantly lower blood calcium levels than non-COVID-19 individuals. [47]. Severe COVID-19 patients had reduced calcium levels [35, 48]. Studies suggest electrolyte level abnormalities in Long-COVID patients, but electrolytes do not serve as severity markers in COVID-19 [49]. Studies have shown that hypermagnesemia significantly indicates the severity and potential adverse outcomes of SARS-CoV-2 infections [35]. COVID-19 patients may experience electrolyte disturbances, including calcium [38]. Our findings are that electrolyte disturbances may be caused by several variables, including the severity of the illness, maintaining calcium and magnesium homeostasis, comorbidities, drugs, or therapy approaches, as a result of the complex physiological imbalances linked to Long-COVID.

Our analysis found that the three Long-COVID biomarkers—Gal, GalR1, and Gal/GalR1—explain a lot of the variance in identifying Long-COVID patients. Figure 1 shows serum Gal (pg/ml) values for Long-COVID patients and healthy controls. Galanin was observed to be considerably higher in Long-COVID patients than in controls. As observed, influenza increases galanin expression in the respiratory tract, which may impact inflammation and local immunological responses [50, 51]. Many inflammatory disorders are linked to Galanin, and other regulatory peptides modify immune system responses [52]. Age-related chronic disorders are linked to the Galanin system [53]. Observe Gal/GalRs ratio system targets in pain, depression, epilepsy, neurodegenerative disorders, diabetes, and cancer [54]. Galanin affects smooth muscle, affecting gastrointestinal and respiratory function [55]. In mammals (guinea pigs), galanin’s distribution implies it may affect airway, vascular, and secretory processes [56]. Galanin analogs may lower post-ictal respiratory collapse [57]. Glanin affects depression, anxiety, pain threshold, and behavior [26]. Galanin reduces acetylcholine release, hence a receptor antagonist may improve cognition [58]. The findings of our study suggest that there may be a correlation between increased levels of Galanin and the persistence or advancement of Long-COVID symptoms, indicating a potential involvement of Galanin in the underlying pathogenesis. Long-COVID patients have elevated GALR1 levels. There is a result that galanin and galanin-like peptides activate GALR1, a galanin receptor [110]. Activating GalR1 inhibits proliferation [59]. GalR1 activates certain galanin receptor subtypes to protect neurons from excitotoxic damage [60]. GALR1 is amplified in nerve tissues [61]. The profile of potency for the regulation of cAMP by cloned GAL1 receptors [62]. Galanin receptors (GALRs) mediate differential signal pathways, and GAL1R activation has antiproliferative effects, suggesting they may be therapeutic targets and diagnostic markers for particular tumors [63]. GALR1 levels may modulate locus coeruleus (LC) signaling and neuroplasticity [64]. It appears that higher GALR1 levels are linked to persistent COVID symptoms. Numerous factors may raise GALR1 levels: Long-COVID or other health conditions may improve GALR1 function. Disease progress or challenge may have boosted galanin signaling. Alterations in GalR1 may affect immunity, pain, and inflammation. Researchers’ methodologies, setting, and conditions may explain GALR1’s growth. Receptor expression variations’ effects need more study. Figure 3 shows Long-COVID patients and healthy controls (HC) Gal/GalR1 ratios. Long-COVID patients exhibited far higher Gal/GalR1 ratios than healthy controls. Consistent results: The system of galanin/galanin-receptors may be able to help discover treatment targets for a variety of illnesses, including pain, mental illnesses, epilepsy, neurodegenerative diseases, diabetes, and cancer [54]. GAL/GalR1 act as tumor suppressors [65]. Lower GAL/GalR1 levels may contribute to gastric cancer growth [29]. Several studies have linked GAL/GALR expressions to tumor differentiation [66]. The observed increase in Galanin levels compared to GALR1 levels was linked to severe symptoms, suggesting a possible dysregulation in the galaninergic system, affecting inflammation, pain modulation, and immune responses. Gal/GALR1 ratios indicate Galanin abundance compared to its receptor, which is high in severe Long-COVID patients. Table 3 shows the Long-COVID group’s biomarker-sociodemographic connection. Significant age-related relationships were found for Galanin and the GAL/GALR1 ratio. SpO2 was linked to Galanin and GALR1. Table 4 illustrates the biomarker associated with cation levels in the LC group. No biomarker-cation association was found. Electrolyte levels may not impact Galanin regulation. It appears that these levels’ homeostasis may not directly alter system regulation. Galanin was correlated with GalR1 and Gal/GalR1 and, respectively. GALR1 was associated with Gal/GalR1. The results showed a hydrophobic interaction between galanin and GalR1. It’s unlike other G protein-coupled receptors, which need their peptide ligands to interact with water close or far from the plasma membrane. They also found that the agonist/GalR1 combination penetrates cells soon after binding [67]. The GalR1 receptor activates only the Gi pathway, while GalR2 activates Go, Gq/G11, and Gi, suggesting that the tissue distribution patterns and signalling profiles of GalR1 and GalR2 may underlie the functional spectra of Galanin action mediated by these receptors and regulate the diverse physiological functions of galanin [68]. The study found Galanin system interactions, with positive associations with GALR1 and the Gal/GalR1 ratio indicating coordinated responses and negative correlations indicating complicated regulatory links.

The diagnostic sensitivity and specificity of Galanin, GALR1, and Gal/GalR1 for LC were determined using ROC analysis curves are shown in Figure 3. As shown in Table 5 and Figure 4, Galanin and GalR1 have the highest sensitivities (71.7% and 60.0%, respectively) for distinguishing Long-COVID patients from healthy controls. Since Galanin and GalR1 are not diagnostic, the table results can be understood as follows: Subjects with Galanin levels over the cut-off value (53.302 pg/ml) may have LC with a sensitivity of 71.7% and a specificity of 73.3%. A GALR1 (pg/ml) increase over the cut-off value (1.815 pg/ml) suggests LC patients with a sensitivity of 60.0% and a specificity of 60.0%. A GalR1/Gal rise beyond the cut-off value (30.74 pg/ml) suggests LC patients with a sensitivity of 58.3% and a specificity of 57.6%. These findings match these studies: The galaninergic system may also affect alcoholism, chronic pain, and bowel and skin inflammatory illnesses [69]. Galanin has a role in experimental autoimmune encephalomyelitis pathogenesis [70]. Sleep-wake control involves neuropeptides like Galanin [71]. Increased Galanin levels were linked to moderate OSAS [72]. Galanin co-release from retrotrapezoid nucleus neurons may counterbalance glutamatergic inputs to respiratory centers to downscale energetically wasteful hyperventilation, contributing to neuroplasticity by decreasing ventilation [73]. GALR1 controls galanergic signaling by acting as a dimer outside a cell and moving inside when triggered by galanin [74]. The Gal/GalR1 peptide receptor regulates afferent ending excitability in chronic tissue injury and pain via an inhibitory presynaptic autocrine action [75]. No association was seen between GAL binding and proliferation [76]. Chronic neuronal inflammation and injury may cause neurological and cognitive deficits since SARS-CoV-2 can breach the blood-brain barrier [77]. ROC analysis showed Galanin and GALR1 biomarkers had the highest sensitivities, suggesting they could distinguish Long-COVID patients from healthy controls. However, the Gal/GalR1 ratio did not show significant diagnostic ability, highlighting the need for appropriate markers.

## Conclusions

It is the first study to find elevated serum galanin and GALR1 levels in COVID patients, particularly those with anosmia and other persistent symptoms. Long-COVID disease correlates to galanin, GALR1, and other neural biomarkers, which can help disease monitoring in Long-COVID patients. Our findings need to be confirmed in a larger population. These observations allow further study of galanin and GALR1 in respiratory infectious illnesses.

### Limitation

The study of long-term COVID patients faces challenges such as diverse symptoms, lack of consensus, population variability, follow-up uncertainties, recall bias, biomarker limitations, ethical concerns, and resource-intensive nature. Correlating IL-6 levels with neuronal biomarkers like galanin is crucial for estimation, improving research outcomes, and understanding Long-COVID.

### Ethics statement

The institutional ethics committee at the University of Kufa (1657/2023) reviewed and authorized the studies using human subjects. The participants/patients gave written informed consent to take part in this investigation.

### Human and animal rights

The study was carried out ethically in accordance with the World Medical Association Declaration of Helsinki and compliance with Iraqi, international, and privacy legislation. Additionally, following the Declaration of Helsinki, the Belmont Report, the CIOMS Guidelines, and the International Conference on Harmonisation in Good Clinical Practice (ICH-GCP), our IRB abides by the International Guidelines for the Protection of Human Research Subjects.

## CONSENT FOR PUBLICATION

Before participating in this study, each subject provided written informed consent.

## Data Availability

All data produced in the present study are available upon reasonable request to the authors

## Acknowledgments

The authors express gratitude to various hospitals in Iraq for their assistance in gathering sample material, senior pulmonologists Maytham Abdulameer Al Maamory and Ammar Abbas Neamh, and internal lab staff for estimating biomarker levels.

